# Checklists to Detect Potential Predatory Biomedical Journals: A Systematic Review

**DOI:** 10.1101/19005728

**Authors:** Samantha Cukier, Lucas Helal, Danielle B Rice, Justina Pupkaite, Nadera Ahmadzai, Mitchell Wilson, Becky Skidmore, Manoj Lalu, David Moher

## Abstract

**Background:** We believe there is a large number of checklists to help authors detect predatory journals. It is uncertain whether these checklists contain similar content.

**Purpose:** Perform a systematic review to identify checklists to detect potential predatory journals and to examine their content and measurement properties.

**Data Sources:** MEDLINE, Embase, PsycINFO, ERIC, Web of Science and Library, Information Science & Technology Abstracts (January 2012 to November 2018), university library websites (January 2019), YouTube (January 2019).

**Study Selection:** Original checklists used to detect potential predatory journals published in English, French or Portuguese, with instructions in point form, bullet form, tabular format or listed items, not including lists or guidance on recognizing “legitimate” or “trustworthy” journals.

**Data Extraction:** Pairs of reviewers independently extracted study data and assessed checklist quality and a third reviewer resolved conflicts.

**Data Synthesis:** Of 1528 records screened, 93 met our inclusion criteria. The majority of included checklists were in English (n = 90, 97%), could be completed in fewer than five minutes (n = 68, 73%), had an average of 11 items, which were not weighted (n = 91, 98%), did not include qualitative guidance (n = 78, 84%) or quantitative guidance (n = 91, 98%), were not evidence-based (n = 90, 97%) and covered a mean of four (of six) thematic categories. Only three met our criteria for being evidence-based.

**Limitations:** Limited languages and years of publication, searching other media.

**Conclusions:** There is a plethora of published checklists that may overwhelm authors looking to efficiently guard against publishing in predatory journals. The similarity in checklists could lead to the creation of evidence-based tools serving authors from all disciplines.

**Funding Source:** This project received no specific funding. David Moher is supported by a University Research Chair (University of Ottawa). Danielle Rice is supported by a Canadian Institutes of Health Research Health Systems Impact Fellowship; Lucas Helal is supported by Coordenação de Aperfeiçoamento de Pessoal de Nível Superior - Brasil (CAPES) - Finance Code 001, PDSE - 88881.189100/2018 - 01. Manoj Lalu is supported by The Ottawa Hospital Anesthesia Alternate Funds Association.

## INTRODUCTION

The influx of predatory publishing along with the substantial increase in the number of predatory journals, pose a risk to scholarly communication (1,2). Predatory journals often lack an appropriate peer-review process and frequently are not indexed (3), yet authors are required to pay an article processing charge. The lack of quality control, the inability to effectively disseminate research and the lack of transparency compromise the trustworthiness of articles published in these journals. Currently, no definition of predatory journals exists, however, characteristics have been studied (3) and a consensus definition is currently being developed (4). Lists of suspected predatory journals and publishers are available, although different criteria for inclusion are used (5).

To help prospective authors avoid predatory journals, various groups have developed checklists. Armed with this knowledge, it is hoped that there will be a substantial decrease in submissions to these journals. Anecdotally, we have recently noticed a steep rise in the number of checklists developed for this purpose, although this has not previously been quantified. Large numbers of checklists with varying content may confuse authors, and possibly make it more difficult for them to choose any one checklist, if any at all, as suggested by the *choice overload* hypothesis (6). The net effect of the abundance of conflicting information could be that users will not consult any checklist and these efforts will be lost, or discrepancies between checklists could impact the credibility of each one. We performed a systematic review of the content and measurement properties of checklists used to detect probable predatory journals.

## METHODS

We followed standard procedures for systematic reviews and reported results according to Preferred Reporting Items for Systematic reviews and Meta-Analyses (PRISMA) guidelines (7). The project protocol was publicly posted prior to data extraction on the Open Science Framework (http://osf.io/g57tf).

### Data Sources and Searches

An experienced medical information specialist (BS) developed and tested the search strategy using an iterative process in consultation with the review team. The strategy was peer reviewed by another senior information specialist prior to execution using the PRESS Checklist (8) (see Appendix 1).

We searched multiple databases with no language restrictions. Using the OVID platform, we searched Ovid MEDLINE® ALL (including in-process and epub-ahead-of-print records), Embase Classic + Embase, PsycINFO and ERIC. We also searched Web of Science and the Library, Information Science and Technology Abstracts (LISTA) database (Ebsco platform). The LISTA search was performed on November 16, 2018 and the Ovid and Web of Science searches were performed on November 19, 2018. Retrieval was limited to the publication dates 2012 to the present. We used 2012 as a cut-off since data about predatory journals were first collected in 2010 (9), and became part of public discourse in 2012 (10). The search strategy for the Ovid databases is included in Appendix 2.

We searched two relevant sources of unpublished grey literature for checklists that help identify predatory journals and/or publishers: 1) University library websites of the top 10 universities in each of the four world regions (Americas, Europe, Asia / Oceania, Africa), as identified by the Shanghai Academic Ranking of World Universities (http://www.shanghairanking.com/ARWU-Statistics-2018.html) and the library websites of Canada’s most research-intensive universities (U15) (search date January 18, 2019); and 2) YouTube videos that contained checklists (search date January 6, 2019). We limited our YouTube search to the top 50 results filtered by “relevance” and used a private browser window. Detailed methods of these searches are available on the Open Science Framework (http://osf.io/g57tf.).

### Eligibility criteria

#### Inclusion criteria

We included studies and/or original checklists developed or published in English, French or Portuguese (languages spoken by the authors). We defined checklist as a tool whose purpose is to detect a potential predatory journal where the instructions are in point form / bullet form / tabular format / listed items. To be an original checklist, items had to have been identified and/or developed by the study authors or include a novel combination of items from multiple sources, or an adaptation of another checklist plus items added by the study authors. We first included studies that discussed the use of an already existing checklist; we used these studies to search out the paper that discussed the original checklist development and then included only the study that discussed the original checklist development.

#### Exclusion criteria

Checklists were not considered original if items were hand-picked from other sources; for example, if authors identified the five most salient points from an already existing checklist. We did not include lists or guidance on recognizing a “legitimate” or “trustworthy” journal.

### Study selection

Following de-duplication of the identified titles, we screened records using the online systematic review software program Distiller Systematic Review (DSR) (Evidence Partners Inc., Ottawa, Canada). For each stage of screening, data extraction and risk of bias assessment, we pilot tested a 10% sample of records among five to six reviewers. Screening was performed in two stages. First (stage 1), based on title and abstract and second (stage 2), based on full-text screening, was administered by two reviewers independently and in duplicate. At both stages, discrepancies were resolved through discussion between the reviewers and consultation with a third reviewer, where necessary.

### Data Extraction and Risk of Bias Assessment

For each eligible study, two reviewers independently extracted relevant data into DSR and a third reviewer resolved any conflicts. The extracted data items were as follows: Checklist name, number of items in the checklist, whether or not the items were weighted, the number of thematic categories covered by the checklist (six-item list developed by Cobey et al. (3)), publication details (name of publication, author and date of publication), approximate time to complete checklist (reviewers used a timer to emulate the process that a user would go through to use the checklist and recorded the time as 0-5 minutes, 6-10 minutes, or more than 10 minutes), language of the checklist, whether or not the checklist was translated and into what language(s), methods used to develop the checklist (details on data collection, if any), whether or not there was qualitative guidance (instructions on how to use the checklist and what to do with the results) and/or quantitative guidance (instructions on summing the results or quantitatively assessing the results to inform a decision). The list of extracted data items can be found on the Open Science Framework (https://osf.io/na756/).

In assessing checklists identified via YouTube, we extracted data items that were presented visually so that any item or explanation that was delivered by audio only was not included in our assessment. For example, if presenters only talked about a checklist item but did not have it on a slide in the video or in a format that could be seen by those watching the video, we did not extract this data.

To assess risk of bias, we created an a priori list of five questions developed by the research team for the purpose of this review, adapted from A Checklist for Checklists tool (11) and principles of internal and external validity (12). The creation of a novel tool to assess risk of bias was necessary since no formal assessment tool exists for the type of review we conducted. We used the results of this assessment to determine whether or not the checklist was evidence-based. We assigned each of the five criterion (listed below) a judgement of “yes” (i.e. low risk of bias), “no” (i.e. high risk of bias) or “can?t tell” (i.e. unclear risk of bias) (see coding manual with instructions for assessment to determine risk of bias ratings: https://osf.io/sp4vx/). If the checklist scored three or more “yes” answers on the questions below, we considered it evidence-based. Two reviewers independently assessed data quality in DSR where discrepancies were resolved through discussion. A third reviewer was called to resolve conflicts where necessary.

The five criteria used to assess risk of bias in this review were as follows:

1. Did the developers of the checklist represent more than one stakeholder group (e.g. researchers, academic librarians, publishers)?
2. Do the developers report gathering any data for the creation of the checklist (i.e. conduct a study on potential predatory journals, carry out a systematic review, collect anecdotal data)?
3. Does the checklist meet at least one of the following criteria: (1) Has title that reflects its objectives; (2) Fits on one page; (3) Each item on the checklist is one sentence?
4. Was the checklist pilot-tested or trialed with front-line users (e.g. researchers, students, academic librarians)?
5. Do the authors report how many criteria in the checklist a journal must meet in order to be considered predatory?

In assessing websites, we used a “two-click rule” to locate information. Once on the checklist website, if we did not find the information within two mouse clicks, we determined no information was available.

### Data Synthesis and Analysis

We examined the checklists qualitatively and conducted qualitative comparisons of the items between the included checklists to gauge their agreement on content by item and overall. We summarized checklists in table format to facilitate inspection and discussion of findings. Frequencies and percentages were used to present characteristics of the checklists. We used the checklist developed by Shamseer et al. (13) as the reference checklist and compared our results to the reference list. We chose this as the reference list because of the rigorous empirical data provided by authors to ascertain characteristics of potential predatory journals.

## RESULTS

### Deviations from our protocol

We refined our definition of an *original* checklist to exclude checklists that were comprised of items taken solely from another checklist. Checklists made up of items taken from more than one source were considered original even when the developers did not create the checklist themselves. For reasons of feasibility, we did not search the reference lists of included studies to identify further potentially relevant studies.

We had anticipated using the liberal accelerated method to screen titles and abstracts. Instead, we had two reviewers screen records independently and in duplicate. We changed methods because it became feasible to use the traditional screening approach, which also reduced the required number of full-text articles to be ordered.

After completing data collection, we recognized that checklists were being published in discipline-specific journals. We wanted to determine what disciplines were represented and in what proportion. We conducted a scan of the journals and used an evolving list of disciplines to assign to the list of journals.

### Study selection

Following the screening of 1433 records, we identified 93 original checklists to be included in our study, made up of 53 records from an electronic database search, 30 from university library websites and 10 from YouTube (see full details in Figure 1: PRISMA flow diagram).

### Study characteristics

We identified 53 checklists identified through our search of electronic databases. The numbers of checklists identified increased over time: one each in 2012 (10), 2013 (14) and then increases in the most recent years (16 in 2017 (13,15–29) and 12 in 2018 (30–41)). We identified 30 original checklists (1,42–70) from university library websites. More checklists were published in more recent years (2017 = 4 (44–47); 2018 = 7 (48–54); 2019 = 11 (55–65); five checklists listed no publication date). We identified 10 more checklists from YouTube (71–80) that included one uploaded in 2015 (71), six in 2017 (72–77) and three in 2018 (78–80). See Table 1 for full study characteristics.

**Table 1.** Characteristics of checklists (oldest to most recently published)

| Study | Checklist name | Number of items | Items weighted Y/N | Time to complete min | Methods used to develop checklist (NR = Not reported) | Qualitative guidance Y/N | Quantitative guidance Y/N |
| --- | --- | --- | --- | --- | --- | --- | --- |
| **Checklists from electronic journal databases n = 53** |  |  |  |  |  |  |  |
| Index checklist Shamseer, 2017 (13) | Salient characteristics of potential predatory journals | 13 | N | 0-5 | Cross-sectional analysis 93 predatory journals, 99 OA, 100 subscription based journals assessed | No | No |
| Beall, 2012 (10) | Criteria for Determining Predatory Open-Access Publishers | 5 | No | 10+ min | Criteria based on AOSPA<sup>1</sup> Code of Conduct, two COPE<sup>2</sup> publications | No | No |
| Beall, 2013 (14) | Some warning signs of questionable publishers | 7 | No | 0-5 min | Observational research on own emails received | No | No |
| Crawford, 2014 (100) | No title | 11 | No | 6-10 min | Assessed all criteria in Beall's criteria | Yes | No |
| Knoll, 2014 (81) | Avoiding Predatory OA Journals | 17 | No | 0-5 min | Works cited | No | No |
| Lukic, 2014 (82) | No title | 13 | No | 0-5 min | Multiple references | No | No |
| Beall, 2015 (102) | Criteria for Determining Predatory Open-Access Publishers | 5 | No | 10+ min | Criteria based on COPE documents: Code of Conduct and Principles of Transparency and Best Practices in Scholarly Publication | No | No |
| Bhad, 2015 (83) | How should one suspect a journal could be a predatory journal? | 9 | No | 0-5 min | NR | No | No |
| Bradley-Springer, 2015 (84) | No title | 6 | No | 0-5 min | Multiple references | No | No |
| Hemmat Esfe, 2015 (85) | Features of the Fake Journals | 9 | No | 0-5 min | NR | No | No |
| INANE Predatory Publishing Practices Collaborative, 2015 (87) | Guidelines for evaluating the integrity of a journal / Red flags | 7 | No | 0-5 min | Limited literature review | No | No |
| Pamukcu Gunaydin, 2015 (86) | No title | 10 | No | 0-5 min | Authors' top 10 based on other references | No | No |
| Proehl, 2015 (88) | Guidelines for evaluating the integrity of a journal - | 7 | No | 0-5 min | References to other checklists: COPE etc. | No | No |

<sup>1</sup> OASPA = Open Access Scholarly Publishers Association<sup>2</sup> COPE = Committee on Publication Ethics

|  |  |  |  |  |  |  |  |
| --- | --- | --- | --- | --- | --- | --- | --- |
|  | Red flags |  |  |  |  |  |  |
| Stone, 2015 (89) | Guidelines for evaluating the integrity of a journal | 6 | No | 0-5 min | Other credible resources: COPE, INANE<sup>3</sup>, other | No | No |
| Yucha, 2015 (90) | Guidelines for Evaluating the Integrity of a Journal - Red flags | 10 | No | 0-5 min | Multiple references to other checklists | No | No |
| Cariappa, 2016 (91) | Telltale signs - Something is wrong! | 7 | No | 0-5 | Some literature cited | No | No |
| Carroll, 2016 (92) | Common Practices of Predatory Open Access Publications | 4 | No | 0-5 min | Limited literature review | No | No |
| Dadkhah, 2016 (103) | Criteria to rank predatory journals | 14 | Yes | 10+ min | Observational study of 150 journals 80 predatory, 70 non | Yes | Yes |
| Fraser, 2016 (93) | Red Flags for Recognizing Predatory Journals | 6 | No | 0-5 min | Two citations | No | No |
| Glick, 2016 (94) | What you can expect from a predatory publisher | 7 | No | 0-5 min | Multiple citations | No | No |
| Glick, 2016a (95) | Clues suggesting a "predatory" journal | 11 | No | 0-5 min | Multiple references | No | No |
| Hansoti, 2016 (96) | Overall Approach to Choosing the Journal | 11 | No | 0-5 min | Extensive literature review | Yes | No |
| Morley, 2016 (97) | 10 steps to spot a predatory publisher | 10 | No | 0-5 min | A few citations | Yes | No |
| Nolan, 2016 (98) | None section title exists but not title | 5 | No | 0-5 min | None noted | No | No |
| Ward, 2016 (101) | No title | 8 | No | 6-10 min | None listed | No | No |
| Abadi, 2017 (15) | No title | 26 | No | 6-10 min | NR | No | No |
| Balehegn, 2017 (29) | No title | 5 | No | 0-5 min | References | No | No |
| Berger, 2017 (16) | Detailed Characteristics of Predatory Journals | 15 | No | 6-10 min | NR | No | No |
| Das, 2017 (17) | How to identify predators? | 15 | No | 0-5 min | Two citations | No | No |
| Erfanmanesh 2017 (18) | No title | 18 | No | 6-10 min | Multiple references | No | No |
| Eriksson, 2017 (19) | Characteristics of a predatory journal | 25 | No | 10+ | Limited literature review | No | No |
| Janodia, 2017 (20) | No title | 9 | No | 6-10 min | NR | Yes | No |
| Khan, 2017 (21) | Attributes, characteristics and | 9 | No | 0-5 min | Citations | No | No |

<sup>3</sup> INANE = International Academy of Nursing Editors

|  |  |  |  |  |  |  |  |
| --- | --- | --- | --- | --- | --- | --- | --- |
|  | practices of potential predatory journals |  |  |  |  |  |  |
| Klyce, 2017 (22) | Common characteristics of predatory journals | 13 | No | 0-5 min | Limited literature review | No | No |
| Manca, 2017 (23) | No title | 6 | No | 0-5 min | Limited literature review | No | No |
| Miller, 2017 (24) | Signs of a Predatory Publisher | 8 | No | 0-5 min | NR | No | No |
| Misra, 2017 (25) | Red flags based on which one may suspect the legitimacy of a journal | 17 | No | 0-5 min | Literature review and authors' experiences | No | No |
| Mouton, 2017 (26) | Comparing the characteristics of good practice in scholarly publishing with those of predatory publishing | 7 | No | 0-5 min | In-depth assessment of journals identified by Beall's list where South African authors published | No | No |
| Oren, 2017 (27) | Obvious signs of predatory journals | 7 | No | 0-5 min | NR | No | No |
| Shamseer, 2017 (13) | Salient characteristics of potential predatory journals | 13 | No | 0-5 min | Observational study 93 predatory journals, 99 OA<sup>4</sup>, 100 subscription-based journals assessed | No | No |
| Stratton, 2017 (28) | Characteristics of Health and Medical Journal Publishing Formats - Open Access Predatory | 4 | No | 0-5 min | Cited references | No | No |
| Ajuwon, 2018 (32) | Characteristics of Predatory Publishers and Journals | 12 | No | 0-5 min | Citations from other sources | No | No |
| Bowman, 2018 (33) | Identifying Predatory Journals and Publishers | 29 | No | 6-10 min | NR | No | No |
| Gerberi, 2018 (34) | Quick List of Predatory Publisher Warning Signs | 7 | No | 0-5 min | Limited literature review | No | No |
| Kokol, 2018 (36) | No title | 4 | No | 0-5 | Analysis of papers 2013-2017 predatory Beall's vs non | No | No |
| Lewinski, 2018 (37) | Eight tips to identify a predatory journal or publisher | 8 | No | 0-5 | NR | Yes | No |
| McCann, 2018 (30) | Guidelines for authors to avoid predatory | 25 | No | 0-5 min | Brief literature review | Yes | No |

<sup>4</sup> OA = Open Access

|  | publishers |  |  |  |  |  |  |
| --- | --- | --- | --- | --- | --- | --- | --- |
| Memon, 2018 (38) | No title | 14 | No | 6-10 min | Collecting emails and web pages of each journal /publisher. Used Beall's list, PubMed, DOAJ<sup>5</sup>, Thomson and Reuters now Clarivate Analytics | No | No |
| Nnaji, 2018 (39) | No title | 11 | No | 6-10 min | Two references | No | No |
| O'Donnell, 2018 (31) | Identifying a predator | 17 | No | 6-10 min | Other evidence-based checklist | No | No |
| Pamukcu Gunaydin, 2018 (35) | How to avoid sending your work to a predatory journal | 5 | No | 0-5 | Limited literature review | No | No |
| Power, 2018 (40) | No title | 11 | No | 6-10 min | References COPE, INANE | No | No |
| Richtig, 2018 (41) | Criteria identified or suggested in the literature that can potentially be used to identify predatory journals | 13 | No | 6-10 | Literature review | No | No |
| Wikipedia, 2019 (99) | No title | 10 | No | 0-5 min | Multiple citations | No | No |
| Study | Checklist name | Number of items | Items weighted Y/N | Time to complete min | Methods used to develop checklist | Qualitative guidance Y/N | Quantitative guidance Y/N |
| **Checklists from university library websites n = 30** |  |  |  |  |  |  |  |
| [REFERENCE CHECKLIST] Shamseer, 2017 | Salient characteristics of potential predatory journals | 13 | N | 0-5 | Cross sectional analysis 93 predatory journals, 99 OA, 100 subscription-based journals assessed | No | No |
| Carlson, 2014 (42) | None | 11 | No | 0-5 min | Based on personal experiences looking into questionable OA journals | No | No |
| Clark, 2015 (1) | None | 5 | No | 0-5 min | NR | No | No |
| University of Edinburgh, 2015 (43) | Some warning signs to look out for | 4 | No | 0-5 min | NR | No | No |
| Africa Check, 2017 (44) | None | 7 | No | 0-5 min | NR | No | No |
| Cabell's – Clarivate, 2017 (45) | None | 64 | No | 10+ min | Analysis by specialists see: https://www2.cabells.com/ | No | No |

<sup>5</sup> DOAJ = Directory of Open Access Journals

|  |  |  |  |  |  |  |  |
| --- | --- | --- | --- | --- | --- | --- | --- |
|  |  |  |  |  | about-blacklist |  |  |
| Duke University Medical Center, 2017 (46) | Be iNFORMEd Checklist | 6 | No | 0-5 min | NR | Yes | No |
| University of Calgary, 2017 (47) | No title | 6 | No | 0-5 min | NR | No | No |
| Coopérer en information scientifique et technique, 2018 (48) | No title | 35 | No | 10+ min | NR | No | No |
| Eaton, University of Calgary 2018 (49) | No title | 12 | No | 0-5 min | Other sources cited | Yes | No |
| Lapinski, Harvard, 2018 (50) | No title | 3 | No | 0-5 min | NR | Yes | No |
| Sorbonne Université, 2018 (51) | Comment repérer un éditeur prédateur | 12 | No | 0-5 min | NR | No | No |
| University of Alberta, 2018 (53) | No title | 19 | No | 6-10 min | NR | No | No |
| University of British Columbia, 2018 (52) | No title | 16 | No | 6-10 min | NR | No | No |
| University of Toronto Libraries, 2018 (54) | Identifying Deceptive Publishers: A Checklist | 22 | Yes | 6-10 min | NR | Yes | Yes |
| Dalhousie University, 2019 (55) | How to recognize predatory journals | 6 | No | 0-5 min | NR | No | No |
| McGill University, 2019 (56) | No title | 4 | No | 0-5 min | NR | No | No |
| McMaster University, 2019 (57) | No title | 6 | No | 0-5 min | NR | No | No |
| Prater, American Journal Experts, 2019 (58) | No title | 8 | No | 0-5 min | NR | Yes | No |
| Ryerson University Library, 2019 (59) | No title | 5 | No | 0-5 min | NR | No | No |
| Université Laval, 2019 (60) | No title | 8 | No | 0-5 min | NR | No | No |
| University of Cambridge, 2019 (61) | No title | 9 | No | 6-10 min | NR | No | No |
| University of Pretoria, 2019 (62) | No title | 19 | No | 0-5 min | NR | No | No |
| University of Queensland Library, 2019 (64) | No title | 6 | No | 0-5 min | NR | No | No |
| University of Queensland Library, 2019a (63) | Red Flags for Open Access Journals | 9 | No | 0-5 min | NR | Yes | No |
| University of Witwatersrand, 2019 (65) | Predatory Publisher Checklist | 26 | No | 10+ min | NR | No | No |
| Canadian Association of Research Libraries, ND (66) | How to assess a journal A.K.A. How not to publish in an undesirable journal | 12 | No | 0-5 min | NR | Yes | No |
| Columbia University | No title | 5 | No | 0-5 min | NR | Yes | No |

| Libraries, ND (67) |  |  |  |  |  |  |  |
| --- | --- | --- | --- | --- | --- | --- | --- |
| Technion Library, ND (68) | No title | 10 | No | 0-5 min | NR | No | No |
| UC Berkley, ND (69) | No title | 3 | No | 0-5 min | Cited 1 paper | No | No |
| University of Ottawa<br>Scholarly Communication,<br>ND (70) | No title | 12 | No | 0-5 min | NR | No | No |
| Study | Checklist name | Number<br>of items | Items<br>weighted<br>Y/N | Time to<br>complete<br>min | Methods used to<br>develop checklist | Qualitative<br>guidance<br>Y/N | Quantitative<br>guidance<br>Y/N |
| **Checklists from YouTube n = 10** |  |  |  |  |  |  |  |
| [REFERENCE CHECKLIST]<br>Shamseer, 2017 | Salient characteristics of<br>potential predatory<br>journals | 13 | N | 0-5 | Cross sectional analysis<br>93 predatory journals, 99<br>OA, 100 subscription-based journals assessed | No | No |
| Robbins, Western Sydney<br>University, 2015 (71) | Red Flags | 9 | No | 6-10 min | NR | No | No |
| Attia, 2017 (72) | Spot Predatory<br>Publishers | 4 | No | 0-5 min | NR | No | No |
| Kysh, USC Keck School of<br>Medicine, 2017 (73) | Characteristics of<br>Predatory Publishers | 9 | No | 0-5 min | NR | No | No |
| McKenna, Rhodes<br>University, 2017 (74) | Predatory Publications:<br>Shark Spotting | 7 | No | 0-5 min | NR | No | No |
| Nicholson, University of<br>Witwatersrand, 2017 (75) | Cautionary Checklist | 36 | No | 10+ min | NR | No | No |
| Raszewski, 2017 (76) | What to watch out for | 4 | No | 0-5 min | NR | No | No |
| Seal-Roberts, Springer<br>Healthcare, 2017 (77) | So how do we recognize<br>a predatory publisher? | 10 | No | 0-5 min | NR | No | No |
| Menon, SCMS Group of<br>Educational Institutions,<br>India and Berryman,<br>Cabell's, 2018 (78) | Characteristics of<br>Predatory Journals | 7 | No | 0-5 min | NR | No | No |
| Office of Scholarly<br>Communication, Cambridge<br>University, 2018 (79) | None | 12 | No | 0-5 min | NR | No | No |
| Weigand, UNC Libraries,<br>2018 (80) | No title | 5 | No | 0-5 min | NR | No | No |

#### Language and translation

Almost all checklists were published in English (n = 90, 97%), and the remaining checklists in French (n = 3, 10%) (48,51,60). Two additional English checklists identified through university library websites were translated into French (54,66) and one was translated into Hebrew (68).

#### Approximate time for user to complete checklist, number of items per checklist, and weighting

Most checklists could be completed within five minutes (n= 68, 73%); 17 checklists (18%) could be completed in six to 10 minutes (15,16,18,20,31,33,38–41,52–54,61,71,100,101) and eight checklists (9%) took more than 10 minutes to complete (10,19,45,48,65,75,102,103). Checklists contained a mean of 11 items each, and a range of between three and 64 items. Items were weighted in two checklists (54,103).

#### Qualitative and quantitative guidance

Qualitative guidance on how to use the results of checklists was provided on 15 checklists (16%) (20,30,37,46,49,50,54,58,64,66,67,96,97,100,103), and quantitative guidance was provided on two checklists (54,103), i.e. prescribing a set number of criteria that would identify the journal or publisher as predatory.

#### Methods used to develop checklists

In order to develop the checklists, authors noted using analysis by specialists (45), information from already existing checklists (88,90,102), using existing literature on predatory journals to pick the most salient features to create a new checklist (30,41,96), developing checklists after empirical study (13,26,38,103) or after personal experiences (14).

### Risk of bias assessment

Among all 93 checklists, there were three (3%) assessed as evidence-based (26,96,103) (see Table 2 for risk of bias assessment detailed results including whether or not a checklist was determined to be evidence-based, i.e. rated as low risk of bias for at least three of the criteria).

**Table 2.** Risk of bias assessment. Three ‘Yes’ assessments results in an overall assessment of evidence based.

| Study | Represent 1+ stakeholder groups (Y/N/U)* | Gather data for checklist development (Y/N/Only citations/ U) | Does the checklist meet at least one of these criteria?<br>(count only the last column in total) |  |  |  | Pilot test (Y/N/U) | Includes number of criteria to be considered predatory (Y/N) | Overall assessment (is it evidence-based?) (Y/N) |
| --- | --- | --- | --- | --- | --- | --- | --- | --- | --- |
|  |  |  | Title (Y/N) | Fits on one page (Y/N) | Each item one sentence (Y/N) | Meets at least one of these (Y/N) |  |  |  |
| Checklists from electronic journal databases (n = 53) |  |  |  |  |  |  |  |  |  |
| Reference checklist Shamseer, 2017 (13) | U | Y | Y | Y | Y | Y | U | N | N |
| Beall, 2012 (10) | U | U | N | N | N | Y | U | N | N |
| Beall, 2013 (14) | U | Y | Y | Y | Y | Y | U | N | N |
| Crawford, 2014 (100) | U | U | N | Y | Y | Y | U | N | N |
| Knoll, 2014 (81) | U | Only citations | Y | Y | N | Y | U | N | N |
| Lukic, 2014 (82) | U | Only citations | N | Y | N | Y | U | N | N |
| Beall, 2015 (102) | U | Only citations | N | N | N | N | U | N | N |
| Bhad, 2015 (83) | U | U | Y | Y | Y | Y | U | N | N |
| Bradley-Springer, 2015 (84) | U | Only citations | N | Y | N | Y | U | N | N |
| Hemmat Esfe, 2015 (85) | U | Only citations | N | Y | Y | Y | U | N | N |
| INANE Predatory Publishing Practices Collaborative, 2015 (87) | U | U | Y | Y | Y | Y | U | N | N |
| Pamukcu Gunaydin, 2015 (86) | U | Only citations | N | Y | N | Y | U | N | N |
| Proehl, 2015 (88) | U | U | N | Y | Y | Y | U | N | N |
| Stone, 2015 (89) | U | Only citations | N | Y | Y | Y | U | N | N |
| Yucha, 2015 (90) | U | U | Y | Y | Y | Y | U | N | N |
| Cariappa, 2016 (91) | U | Only citations | Y | Y | Y | Y | U | N | N |
| Carroll, 2016 (92) | U | Y | Y | Y | Y | Y | U | N | N |
| Dadkhah, 2016 (103) | U | Y | Y | Y | Y | Y | Y | Y | Y |
| Fraser, 2015 (93) | U | Only citations | Y | Y | Y | Y | U | N | N |
| Glick, 2016 (94) | U | U | Y | Y | Y | Y | U | N | N |
| Glick, 2016a (95) | U | Only citations | Y | Y | Y | Y | U | N | N |
| Hansoti, 2016 (96) | Y | Y | N | Y | N | Y | U | N | Y |
| Morley, 2016 (97) | U | U | N | N | N | N | U | N | N |
| Nolan, 2016 (98) | U | U | N | Y | N | Y | U | N | N |
| Ward, 2016 (101) | U | U | N | N | N | N | U | N | N |
| Abadi, 2017 (15) | U | U | N | Y | Y | Y | U | N | N |
| Balehegn, 2017 (29) | U | Only citations | N | Y | Y | Y | U | N | N |
| Berger, 2017 (16) | U | U | N | Y | N | Y | U | N | N |
| Das, 2017 (17) | U | Only citations | Y | Y | Y | Y | U | N | N |
| Erfanmanesh, 2017 (18) | U | Only citations | Y | Y | Y | Y | U | N | N |
| Eriksson, 2017 (19) | U | U | Y | Y | Y | Y | U | N | N |
| Janodia, 2017 (20) | U | U | Y | Y | N | Y | U | N | N |
| Khan, 2017 (21) | Y | Only citations | Y | Y | N | Y | U | N | N |
| Klyce, 2017 (22) | U | Only citations | Y | Y | Y | Y | U | N | N |
| Manca, 2017 (23) | U | Only citations | N | Y | N | Y | U | N | N |
| Miller, 2017 (24) | U | U | Y | Y | Y | Y | U | N | N |
| Misra, 2017 (25) | U | Y | Y | Y | Y | Y | U | N | N |
| Mouton, 2017 (26) | U | U | Y | Y | N | Y | Y | Y | Y |
| Oren, 2017 (27) | U | U | Y | Y | Y | Y | U | N | N |
| Shamseer, 2017 (13) | U | Y | Y | Y | Y | Y | U | N | N |
| Stratton, 2017 (28) | U | U | Y | Y | Y | Y | U | N | N |
| Ajuwon, 2018 (32) | U | Only citations | Y | Y | Y | Y | U | N | N |
| Bowman, 2018 (33) | U | U | Y | Y | N | Y | U | N | N |
| Gerberi, 2018 (34) | U | Only citations | Y | Y | N | Y | U | N | N |
| Pamukcu Gunaydin, 2018 (35) | U | Only citations | Y | Y | N | Y | U | N | N |
| Kokol, 2018 (36) | U | Y | N | Y | Y | Y | U | N | N |
| Lewinski, 2018 (37) | U | Y | Y | Y | N | Y | U | N | N |
| McCann, 2018 (30) | U | Y | Y | Y | Y | Y | U | N | N |
| Memon, 2018 (38) | U | Y | Y | Y | Y | Y | U | N | N |
| Nnaji, 2018 (39) | U | Only citations | Y | N | N | Y | U | N | N |
| O'Donnell, 2018 (31) | U | Only citations | N | N | N | N | U | N | N |
| Power, 2018 (40) | U | Only citations | N | N | N | N | U | N | N |
| Richtig, 2018 (41) | U | Only citations | Y | Y | N | Y | U | N | N |
| Wikipedia, 2019 (99) | U | Only citations | N | Y | Y | Y | U | N | N |
|  |  |  | Title (Y/N) | Fits on one page (Y/N) | Each item one sentence (Y/N) | Meets at least one of these (Y/N) |  |  |  |
| Checklists from university library websites (n = 30) |  |  |  |  |  |  |  |  |  |
| Reference checklist Shamseer, 2017 (13) | U | Y | Y | Y | Y | Y | U | N | N |
| Carlson, 2014 (42) | U | Y | N | N | N | N | U | N | N |
| Clark, 2015 (1) | U | U | N | Y | N | Y | U | N | N |
| University of Edinburgh, 2015 (43) | U | U | Y | Y | Y | Y | U | N | N |
| Africa Check, 2017 (44) | U | Only citations | N | Y | N | Y | U | N | N |
| Cabell's - Clarivate, 2017 (45) | Y | U | N | N | Y | Y | U | N | N |
| Duke University Medical Center, 2017 (46) | U | U | N | N | N | N | U | N | N |
| University of Calgary, 2017 (47) | U | U | N | Y | N | Y | U | N | N |
| Coopérer en information scientifique et technique, 2018 (48) | U | U | N | N | Y | Y | U | N | N |
| Eaton, University of Calgary, 2018 (49) | U | Y | N | Y | Y | Y | U | N | N |
| Lapinski, Harvard University, 2018 (50) | U | U | N | N | N | N | U | N | N |
| Sorbonne Université, 2018 (51) | U | U | Y | Y | Y | Y | U | N | N |
| University of Alberta, 2018 (53) | U | U | N | N | N | N | U | N | N |
| University of British Columbia, 2018 (52) | U | Only citations | Y | N | N | Y | U | N | N |
| University of Toronto Libraries, 2018 (54) | Y | U | N | N | N | Y | U | Y | N |
| Dalhousie University, 2019 (55) | U | U | Y | Y | N | Y | U | N | N |
| McGill University, 2019 (56) | U | U | N | Y | N | Y | U | N | N |
| McMaster University, 2019 (57) | U | U | N | Y | Y | Y | U | N | N |
| Prater - American Journal Experts, 2019 (58) | Y | U | N | N | N | N | U | N | N |
| Ryerson University Library, 2019 (59) | U | U | N | Y | N | Y | U | N | N |
| Université Laval, 2019 (60) | U | U | N | Y | N | Y | U | N | N |
| University of Cambridge, 2019 (61) | U | U | N | Y | N | Y | U | N | N |
| University of Pretoria, 2019 (62) | U | U | N | Y | Y | Y | U | N | N |
| University of Queensland Library, 2019 (64) | U | U | N | Y | N | Y | U | N | N |
| University of Queensland Library, 2019a (63) | U | U | N | Y | N | Y | U | N | N |
| University of Witwatersrand, 2019 (65) | U | U | Y | N | Y | Y | U | N | N |
| Canadian Association of Research Libraries, ND (66) | U | U | N | Y | Y | Y | U | N | N |
| Columbia University Libraries, ND (67) | U | U | N | Y | N | Y | U | N | N |
| Technion Library, ND (68) | U | U | N | Y | N | Y | U | N | N |
| UC Berkeley, ND (69) | U | U | N | Y | N | Y | U | N | N |
| University of Ottawa Scholarly Communication, ND (70) | U | U | N | Y | N | Y | U | N | N |
|  |  |  | Title (Y/N) | Fits on one page (Y/N) | Each item one sentence (Y/N) | Meets at least one of these (Y/N) |  |  |  |
| Checklists from YouTube (n = 10) [website URLs available in appendix] |  |  |  |  |  |  |  |  |  |
| Reference checklist Shamseer, 2017 (13) | U | Y (cross-sectional study on journals) | Y | Y | Y | Y | U | N | N |
| Robbins S, Western Sydney University, 2015 (71) | U | U | Y | Y | Y | Y | U | N | N |
| Attia, 2017 (72) | U | U | Y | N | N | Y | U | N | N |
| Kysh, USC Keck School of Medicine, 2017 (73) | U | U | Y | Y | Y | Y | U | N | N |
| McKenna, Rhodes University, 2017 (74) | U | U | Y | Y | Y | Y | U | N | N |
| Nicholson, University of Witwatersrand, 2017 (75) | U | U | Y | N | N | Y | U | N | N |
| Raszewski, 2017 (76) | U | U | Y | Y | N | Y | U | N | N |
| Seal-Roberts, Springer Healthcare, 2017 (77) | U | U | Y | Y | Y | Y | U | N | N |
| Menon, SCMS Group of Educational Institutions, India and Berryman, Cabells, 2018 (78) | U | U | Y | Y | Y | Y | U | N | N |
| Office of Scholarly Communication, Cambridge University, 2018 (79) | U | U | N | N | Y | Y | U | N | N |
| Weigand, UNC Libraries, 2018 (80) | U | U | N | Y | Y | Y | U | N | N |

\*Y = Yes, N = No, U = Unclear

#### Results for risk of bias criteria

##### Criterion #1: Representation of more than one stakeholder group in checklist development

For the majority of checklists (n = 88, 94%), it was unclear whether there was representation of more than one stakeholder group in the checklist development process. The remaining five checklists were developed with representation from more than one stakeholder group (low risk of bias) (21,45,54,58,96).

##### Criterion #2: Authors reported gathering data to inform checklist development

In most studies (n = 55, 59%) there was no mention of data gathering for checklist development (low risk of bias); in 26 cases (28%), one or two citations were noted next to checklist items, with no other explanation of item development or relevance (high risk of bias) (17,18,21–23,29,31,32,34,35,39–41,44,52,81,82,84–86,89,91,93,95,99,102). Twelve records (13%) included a description of authors gathering data to develop a checklist for this criterion (low risk of bias) (13,14,25,30,36–38,42,49,92,96,103).

##### Criterion #3: At least one of the following: Title that reflected checklist objective; Checklist fits on one page; Items were one sentence long

Most checklists were assessed as low risk of bias on this criterion, with 83 of the checklists (90%) meeting at least one of the noted criteria (relevant title, fits on one page, items one sentence long).

##### Criterion #4: Authors reported pilot testing the checklist

In the majority of studies (n = 91, 98%), authors did not report pilot testing during the checklist development stages.

##### Criterion #5: Checklist instructions included a threshold number of criteria to be met in order to be considered predatory

The majority of studies (n = 90, 97%), did not include a threshold number of criteria to be met in order for the journal or publisher to be considered predatory (high risk of bias).

### Assessment of the thematic content of the included checklists

We found checklists covered the six thematic categories, as identified by Cobey et al. (3), as follows (see Table 3 for thematic categories and descriptions of categories):

**Table 3.** Thematic categories covered by the checklists (oldest to most recently published)

| Study | Categories covered by checklist* |  |  |  |  |  |
| --- | --- | --- | --- | --- | --- | --- |
|  | Journal Operations | Article | Editorial and Peer Review | Communications | Article Processing Charge | Dissemination, indexing + archiving |
| **Checklists from electronic journal databases n = 53** |  |  |  |  |  |  |
| [REFERENCE CHECKLIST] Shamseer, 2017 | X | X | X | X | X | X |
| Beall, 2012 (10) | X | X | X | X | X | X |
| Beall, 2013 (14) | X | X | X | X |  |  |
| Crawford, 2014 (100) |  | X | X |  | X |  |
| Knoll, 2014 (81) | X |  | X | X | X | X |
| Lukic, 2014 (82) | X |  | X | X |  | X |
| Beall, 2015 (102) | X | X | X | X | X | X |
| Bhad, 2015 (83) | X |  |  | X | X | X |
| Bradley-Springer, 2015 (84) | X |  |  | X |  |  |
| Hemmat Esfe, 2015 (85) | X |  | X | X | X |  |
| INANE Predatory Publishing Practices Collaborative, 2015 (87) | X |  | X | X |  |  |
| Pamukcu Gunaydin, 2015 (86) | X | X |  | X | X |  |
| Proehl, 2015 (88) | X |  | X | X |  |  |
| Stone, 2015 (89) | X |  | X | X |  |  |
| Yucha, 2015 (90) | X |  | X | X |  |  |
| Cariappa, 2016 (91) | X | X | X | X |  |  |
| Carroll, 2016 (92) | X |  | X | X |  |  |
| Dadkhah, 2016 (103) | X |  | X | X | X | X |
| Fraser, 2016 (93) | X |  | X | X | X |  |
| Glick, 2016 (94) | X |  | X | X | X |  |
| Glick, 2016a (95) |  | X | X | X | X |  |
| Hansoti, 2016 (96) | X | X | X |  |  | X |
| Morley, 2016 (97) | X |  |  | X | X | X |
| Nolan, 2016 (98) | X |  | X |  | X | X |
| Ward, 2016 (101) | X | X | X | X | X | X |
| Abadi, 2017 (15) | X | X | X | X | X | X |
| Balehegn, 2017 (29) |  |  |  |  |  | X |
| Berger, 2017 (16) | X |  | X | X | X | X |
| Das, 2017 (17) | X |  | X |  | X | X |
| Erfanmanesh, 2017 (18) | X | X | X | X | X | X |
| Eriksson, 2017 (19) | X | X | X | X | X | X |
| Janodia, 2017 (20) | X | X |  |  | X |  |
| Khan, 2017 (21) | X |  | X | X | X | X |
| Klyce, 2017 (22) | X | X | X | X |  | X |
| Manca, 2017 (23) | X | X |  |  | X | X |
| Miller, 2017 (24) | X |  | X |  |  | X |
| Misra, 2017 (25) | X | X | X | X | X | X |
| Mouton, 2017 (26) | X |  | X | X |  | X |
| Oren, 2017 (27) | X | X | X | X |  |  |
| Shamseer, 2017 (13) | X | X | X | X | X | X |
| Stratton, 2017 (28) |  |  | X | X | X | X |
| Ajuwon, 2018 (32) | X |  | X | X | X | X |
| Bowman, 2018 (33) | X | X | X | X | X | X |
| Gerberi, 2018 (34) | X | X | X |  |  |  |
| Kokol, 2018 (36) |  | X |  |  |  |  |
| Lewinski, 2018 (37) |  | X | X | X |  | X |
| McCann, 2018 (30) | X |  | X | X | X | X |
| Memon, 2018 (38) | X |  | X | X | X |  |
| Nnaji, 2018 (39) | X | X | X | X | X | X |
| O'Donnell, 2018 (31) | X | X | X | X | X | X |
| Pamukcu Gunyadin, 2018 (35) | X |  | X | X |  | X |
| Power, 2018 (40) | X |  | X | X | X | X |
| Richtig, 2018 (41) | X | X | X | X | X | X |
| Wikipedia 2019 (99) | X |  | X |  | X | X |
| **TOTALS /53 checklists from electronic journal databases (n, %)** | **47, 89** | **24, 45** | **45, 85** | **42, 79** | **34, 64** | **34, 64** |

| Study | Categories covered by checklists* |  |  |  |  |  |
| --- | --- | --- | --- | --- | --- | --- |
|  | Journal Operations | Article | Editorial and Peer Review | Communications | Article Processing Charge | Dissemination, indexing + archiving |
| **Checklists from university library websites n = 30** |  |  |  |  |  |  |
| [REFERENCE CHECKLIST] Shamseer, 2017 (13) | X | X | X | X | X | X |
| Carlson, 2014 (42) | X |  |  | X |  |  |
| Clark, 2015 (1) | X |  | X |  | X | X |
| University of Edinburgh, 2015 (43) | X |  |  | X |  |  |
| Africa Check, 2017 (44) | X |  | X |  |  | X |
| Cabell's – Clarivate, 2017 (45) | X | X | X | X | X | X |
| Duke University Medical Center, 2017 (46) | X | X | X |  | X | X |
| University of Calgary, 2017 (47) | X | X | X |  | X |  |
| Coopérer en information scientifique et technique, 2018 (48) | X |  | X | X | X | X |
| Eaton, University of Calgary, 2018 (49) | X |  | X | X |  | X |
| Lapinski, Harvard, 2018 (50) |  |  |  |  |  | X |
| Sorbonne Université, 2018 (51) | X |  | X | X | X | X |
| University of Alberta, 2018 (53) | X |  |  | X | X | X |
| University of British Columbia, 2018 (52) | X | X | X | X | X | X |
| University of Toronto Libraries, 2018 (54) | X | X | X | X | X | X |
| Dalhousie University, 2019 (55) | X |  |  | X | X | X |
| McGill University, 2019 (56) | X |  | X | X |  | X |
| McMaster University, 2019 (57) | X |  | X | X | X | X |
| Prater, 2019 (58) | X | X | X |  | X |  |
| Ryerson University Library, 2019 (59) | X | X | X | X | X |  |
| Université Laval, 2019 (60) | X |  | X | X | X | X |
| University of Cambridge, 2019 (61) | X | X | X | X | X | X |
| University of Pretoria, 2019 (62) | X |  | X | X | X | X |
| University of Queensland Library, 2019 (64) | X |  | X | X | X | X |
| University of Queensland Library, 2019a (63) | X | X | X |  |  | X |
| University of Witwatersrand, 2019 (65) | X | X | X | X | X | X |
| Canadian Association of Research Libraries, ND | X | X | X | X | X | X |
| (66) |  |  |  |  |  |  |
| Columbia University Libraries, ND (67) | X |  | X |  |  |  |
| Technion Library, ND (68) | X | X |  |  |  | X |
| UC Berkley, ND (69) | X |  |  | X | X | X |
| University of Ottawa Scholarly Communication, ND (70) | X |  | X | X |  | X |
| **Totals /30 checklists from university library websites (n, %)** | **29, 97** | **12, 40** | **23, 77** | **21, 70** | **20, 67** | **24, 80** |
| **Study** | **Categories covered by checklists\*** |  |  |  |  |  |
|  | **Journal Operations** | **Article** | **Editorial and Peer Review** | **Communications** | **Article Processing Charge** | **Dissemination, indexing + archiving** |
| **Checklists from YouTube n = 10** |  |  |  |  |  |  |
| [REFERENCE CHECKLIST] Shamseer, 2017 (13) | X | X | X | X | X | X |
| Robbins S. Western Sydney University, 2015 (71) | X |  | X | X | X |  |
| Attia, 2017 (72) | X |  | X | X |  |  |
| Kysh, USC Keck School of Medicine, 2017 (73) | X |  | X |  | X | X |
| McKenna, Rhodes University, 2017 (74) | X | X |  | X |  |  |
| Nicholson, University of Witwatersrand, 2017 (75) | X | X | X | X | X | X |
| Raszewski 2017 (76) |  |  | X | X | X |  |
| Seal-Roberts, Springer Healthcare, 2017 (77) | X |  | X | X | X |  |
| Menon, SCMS Group of Educational Institutions, India and Berryman, Cabells, 2018 (78) | X |  | X | X | X | X |
| Office of Scholarly Communication, Cambridge University, 2018 (79) | X | X | X |  | X | X |
| Weigand, 2018 UNC Libraries (80) | X |  | X | X |  |  |
| **Totals /10 checklists from YouTube (n, %)** | **9, 90** | **3, 30** | **9, 90** | **8, 80** | **7, 70** | **4, 40** |
| **TOTAL (n, %)** | **85, 91** | **39, 42** | **77, 83** | **71, 93** | **61, 66** | **62, 67** |

\*Categories as described by Cobey et al. 2018 (3), reprinted with permission:
Journal operations: Features related to how the journal conducts its business operations
Article: Features related to articles appearing in the journal
Editorial and peer review: Any aspect of the internal or external review of submitted articles and decisions on what to publish
Communications: How the journal interacts with potential authors, editors, and readers
Article processing charge: Fees taken in by journal as part of their business model
Dissemination, indexing and archiving: Information on how the journal disseminates articles and use of indexing and archiving tools

#### Journal operations

85 checklists (91%) assessed information on the journal?s operations.

#### Assessment of previously published articles

40 checklists (43%) included questions on the quality of articles published in the journal in question.

#### Editorial and peer review process

77 checklists (83%) included questions on the editorial and peer review process.

#### Communication

71 checklists (76%) included an assessment of the manners in which communication is set up between the journal / publisher and the author.

#### Article processing charges

61 checklists (66%) included an assessment of information on article processing charges.

#### Dissemination, indexing and archiving

62 checklists (67%) included suggested ways in which submitting authors should check for information on dissemination, indexing and archiving procedures of the journal and publisher.

Across all 93 checklists, a mean of four out of the six thematic categories was covered, demonstrating similar themes covered by all checklists. Twenty percent of checklists (n = 19), including the reference checklist, covered all six categories (10,13,15,18,19,25,31,33,39,41,45,52,54,61,65,66,75,101,102). Assessment of previously published articles was the category least often included in a checklist (n = 40, 43%), and a mention of the journal operations was the category most often included in a checklist (n = 85, 91%).

### Discipline-specific journals

We observed that there were different checklists published in journals of distinct disciplines, i.e. of the checklists published in academic journals, 10 were published in nursing journals (22%), eight were published in journals related to general medicine (18%), four in emergency medicine journals (9%), four in information science journals (9%), four in psychiatry and behavioural science journals (9%) and the remaining checklists were published in a variety of other discipline-specific journals, with three or fewer checklists per discipline (specialty medicine, paediatric medicine, general medicine and surgery, medical education, dentistry, ecology, and geography).

## DISCUSSION

The number of checklists developed to help identify predatory journals and/or publishers has increased since 2012. Since 2015, the numbers have increased more dramatically (n = 81, 87%). Comparing the 93 checklists we identified to the reference checklist, we observed that on average, the contents of the checklists are similar in terms of items that should be screened, time to complete the checklist, categories or domains covered by the checklist, number of items in the checklist (this number does vary considerably, however the average number of items is more consistent with the reference list) and lack of qualitative and quantitative guidance on completing the checklists. Furthermore, only 3% of checklists were deemed evidence-based, few checklists weighted any items (2%) and few checklists were developed through empirical study (4%).

### Summary of evidence

In total, we identified 93 checklists to help identify predatory journals and/or publishers. A search of electronic databases resulted in 53 original checklists, a search of library websites of top universities resulted in 30 original checklists and a filtered and limited search of YouTube returned 10 original checklists. Overall, checklists could be completed quickly, covered similar categories of topics and were lacking in guidance that would help a user determine if the journal or publisher was indeed predatory.

### Strengths and Limitations

We used a rigorous systematic review process to conduct the study. We also searched multiple data sources including published literature, university library websites, globally, and YouTube. We were limited by the languages of checklists (English, French and Portuguese), however, it has been determined, that the majority of academic literature is published in English (104) and so we are confident we captured the majority of checklists or at least a representative sample. For reasons of feasibility, we were not able to capture all checklists available. Our reference checklist did not qualify as evidence-based when using our predetermined criteria to assess risk of bias; a possible reason could be because the list of characteristics in the reference list was not initially intended as a checklist per se. Creating a useable checklist requires attention to other details, like those we identify in our risk of bias criteria, that perhaps were not attended to by Shamseer et al. because of the difference in its intended purpose.

We noted that the “Think. Check. Submit” checklist (105) was referenced in many publications and we believe it is used often as guidance for authors in order to identify presumed legitimate journals. However, we did not include this checklist in our study because we excluded checklists that help to identify presumed legitimate publications as opposed to potential predatory ones.

## Conclusions

In our search for checklists to help authors identify potential predatory journals, we found great similarity across checklist media and across journal disciplines in which the checklists were published.

Although many of the checklists were published in field-specific journals and / or addressed a specific audience, the content of the lists did not differ much, only a small proportion reported on empirical methods used to design the checklists, and only a few checklists were deemed evidence-based according to our criteria. Importantly, very few offered concrete guidance on using the checklists or offered any threshold that would guide authors to identify definitively if the journal was predatory. The lack of checklists providing threshold values could be due to the fact that a definition of predatory journals does not currently exist (3). The provision of detailed requirements that would qualify a journal as predatory therefore would be a challenge.

With this large number of checklists in circulation, and the lack of explicit and exacting guidelines to identify predatory publications, are authors at continued risk of publishing in journals that do not follow best publication practices? We see some value in discipline-specific lists for the purpose of more effective dissemination. However, this needs to be balanced against the risk of confusing researchers and overloading them with choice (4). If most of the domains in the identified checklists are similar across disciplines, would a single list, relevant in all disciplines, result in less confusion and maximize dissemination and enhance implementation?

In our study, we found no checklist to be optimal. Currently, we would caution against any further development of checklists and instead provide the following as guidance to authors:

Look for a checklist that:

1. Provides a threshold value for criteria to assess potential predatory journals, e.g. if the journal contains these *three* checklist items then we recommend avoiding submission;
2. Has been developed using rigorous evidence, i.e. empirical evidence that is described or referenced in the publication.

Using an evidence-based tool with a clear threshold for identifying potential predatory journals will help reduce the burden of research waste occurring as a result of the proliferation of predatory publications.

## Data Availability

The study protocol, detailed search methods, data extraction and risk of bias methods are available on the Open Science Framework.

http://osf.io/g57tf

## Acknowledgements

The authors would like to thank Raymond Daniel, Library Technician, at the Ottawa Hospital Research Institute, for document retrieval, information support and setup in Distiller.

## Appendix 1

### *PRESS Guideline* 2015— Search Submission & Peer Review Assessment

Reference: McGowan J, Sampson M, Salzwedel DM, Cogo E, Foerster V, Lefebvre C. PRESS Peer Review of Electronic Search Strategies: 2015 guideline statement. *J Clin Epidemiol* 2016;75:40-6. Available: http://www.jclinepi.com/article/S0895-4356(16)00058-5/pdf.

#### Search submission: This section to be filled in by the searcher

Searcher: Becky Skidmore

Date submitted: 7 Nov 2018 Date requested by: 10 Nov 2018 AM

**1. Systematic Review Title**

Systematic Review of Checklists to Detect Potential Predatory Biomedical Journals and Publishers

**2. This search strategy is …**

**2. This search strategy is ...**
|  |  |
| --- | --- |
| X | My PRIMARY (core) database strategy — First time submitting a strategy for search question and database |
|  | My PRIMARY (core) strategy — Follow-up review NOT the first time submitting a strategy for search question and database. If this is a response to peer review, itemize the changes made to the review suggestions |
|  | SECONDARY search strategy— First time submitting a strategy for search question and database |
|  | SECONDARY search strategy — NOT the first time submitting a strategy for search question and database. If this is a response to peer review, itemize the changes made to the review suggestions |

**3. Database** (e.g., MEDLINE, CINAHL) *[mandatory]*

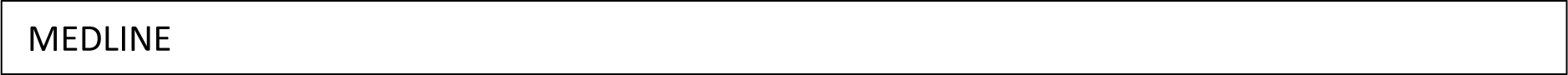

**4. Interface** (e.g., Ovid, EbscoHost…) *[mandatory]*

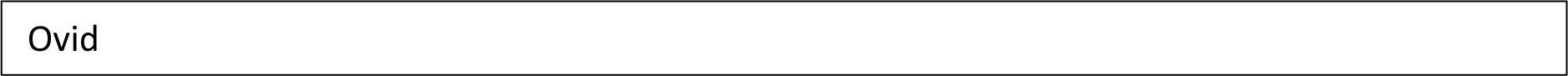

**5. Research Question** (Describe the purpose of the search) *[mandatory]*

**6**.**PICO Format** Outline the PICOs for your question — i.e., <u>P</u>atient, Intervention, <u>C</u>omparison, <u>O</u>utcome, and <u>S</u>tudy Design — as applicable

|  |  |
| --- | --- |
| **P** | Predatory Journals |
| **I / Exposure** | Checklists to detect a potentially predatory journal or publisher |
| **C** |  |
| **O** |  |
| **S** |  |

**7. Inclusion Criteria** (List criteria such as age groups, study designs, etc., to be included) *[optional]*

Publication years 2012-current

**9. Exclusion Criteria** (List criteria such as study designs, date limits, etc., to be excluded) **[optional]**

Opinion pieces and editorials

**10. Was a search filter applied?** No

**If YES, which one(s) (e**.**g**., **Cochrane RCT filter, PubMed Clinical Queries filter)? Provide the source if this is a published filter**. *[mandatory if YES to previous question* — *textbox]*

**11. Notes or comments you feel would be useful for the peer reviewer** *[optional]*

It is possible info will not be restricted to biomedical field so terminology and choice of databases have

been adjusted for this possible broader scope (e.g., also including library science databases, Web of Science)

Have shown Ovid multifile search instead of just MEDLINE

“Series” was suggested as a possible synonym for checklists but was tested and discarded

Much of the vocabulary pertaining to predatory journals has been previously PRESSed (dark and rogue have been added)

There will be an extensive follow-up grey literature search of library web sites, YouTube, etc.

Volume in published literature is very small – one option is to only search on the population (predatory journals) for the electronic database searches (684 records after removing duplicates in Ovid).

Thoughts?

**12. Please copy and paste your search strategy here, exactly as run, including the number of hits per line. [mandatory]**

Database: Embase Classic+Embase <1947 to 2018 November 06>, Ovid MEDLINE(R) ALL <1946 to

November 06, 2018>, PsycINFO <1806 to October Week 5 2018>, ERIC <1965 to August 2018> Search Strategy:

--------------------------------------------------------------------------------

1. (predator* adj3 edit*).tw,kw,kf. (29)
2. (predator* adj3 journal*).tw,kw,kf. (393)
3. (predator* adj3 periodical?).tw,kw,kf. (6)
4. (predator* adj3 publication?).tw,kw,kf. (48)
5. (predator* adj3 publish*).tw,kw,kf. (373)
6. (bogus adj3 edit*).tw,kw,kf. (2)
7. (bogus adj3 journal*).tw,kw,kf. (7)
8. (bogus adj3 periodical?).tw,kw,kf. (0)
9. (bogus adj3 publication?).tw,kw,kf. (0)
10. (bogus adj3 publish*).tw,kw,kf. (1)
11. (dark adj3 edit*).tw,kw,kf. (32)
12. (dark adj3 journal*).tw,kw,kf. (9)
13. (dark adj3 periodical?).tw,kw,kf. (4)
14. (dark adj3 publication?).tw,kw,kf. (2)
15. (dark adj3 publish*).tw,kw,kf. (19)
16. (decepti* adj3 edit*).tw,kw,kf. (21)
17. (decepti* adj3 journal*).tw,kw,kf. (15)
18. (decepti* adj3 periodical?).tw,kw,kf. (0)
19. (decepti* adj3 publication?).tw,kw,kf. (3)
20. (decepti* adj3 publish*).tw,kw,kf. (20)
21. (disreput* adj3 edit*).tw,kw,kf. (0)
22. (disreput* adj3 journal*).tw,kw,kf. (3)
23. (disreput* adj3 periodical?).tw,kw,kf. (0)
24. (disreput* adj3 publication?).tw,kw,kf. (3)
25. (disreput* adj3 publish*).tw,kw,kf. (0)
26. (distrust* adj3 edit*).tw,kw,kf. (1)
27. (distrust* adj3 journal*).tw,kw,kf. (2)
28. (distrust* adj3 periodical?).tw,kw,kf. (0)
29. (distrust* adj3 publication?).tw,kw,kf. (0)
30. (distrust* adj3 publish*).tw,kw,kf. (5)
31. (exploit* adj3 edit*).tw,kw,kf. (107)
32. (exploit* adj3 journal*).tw,kw,kf. (29)
33. (exploit* adj3 periodical?).tw,kw,kf. (1)
34. (exploit* adj3 publication?).tw,kw,kf. (37)
35. (exploit* adj3 publish*).tw,kw,kf. (93)
36. (fake? adj3 edit*).tw,kw,kf. (11)
37. (fake? adj3 journal*).tw,kw,kf. (36)
38. (fake? adj3 periodical?).tw,kw,kf. (0)
39. (fake? adj3 publication?).tw,kw,kf. (4)
40. (fake? adj3 publish*).tw,kw,kf. (20)
41. (hoax$2 adj3 edit*).tw,kw,kf. (1)
42. (hoax$2 adj3 journal*).tw,kw,kf. (5)
43. (hoax$2 adj3 periodical?).tw,kw,kf. (0)
44. (hoax$2 adj3 publication?).tw,kw,kf. (2)
45. (hoax$2 adj3 publish*).tw,kw,kf. (4)
46. (illegitim* adj3 edit*).tw,kw,kf. (3)
47. (illegitim* adj3 journal*).tw,kw,kf. (19)
48. (illegitim* adj3 periodical?).tw,kw,kf. (0)
49. (illegitim* adj3 publication?).tw,kw,kf. (6)
50. (illegitim* adj3 publish*).tw,kw,kf. (12)
51. (mislead* adj3 edit*).tw,kw,kf. (42)
52. (mislead* adj3 journal*).tw,kw,kf. (36)
53. (mislead* adj periodical?).tw,kw,kf. (0)
54. (mislead* adj3 publication?).tw,kw,kf. (57)
55. (mislead* adj publish*).tw,kw,kf. (5)
56. (non-legitim* adj3 edit*).tw,kw,kf. (0)
57. (non-legitim* adj3 journal*).tw,kw,kf. (0)
58. (non-legitim* adj3 periodical?).tw,kw,kf. (0)
59. (non-legitim* adj3 publication?).tw,kw,kf. (0)
60. (non-legitim* adj3 publish*).tw,kw,kf. (0)
61. (questionabl* adj3 edit*).tw,kw,kf. (24)
62. (questionabl* adj3 journal*).tw,kw,kf. (38)
63. (quesionabl* adj3 periodical?).tw,kw,kf. (0)
64. (questionabl* adj3 publication?).tw,kw,kf. (44)
65. (questionabl* adj3 publish*).tw,kw,kf. (48)
66. (racket? adj3 edit*).tw,kw,kf. (0)
67. (racket? adj3 journal*).tw,kw,kf. (1)
68. (racket? adj3 periodical?).tw,kw,kf. (0)
69. (racket? adj3 publication?).tw,kw,kf. (0)
70. (racket? adj3 publish*).tw,kw,kf. (0)
71. (rogue adj3 edit*).tw,kw,kf. (4)
72. (rogue adj3 journal*).tw,kw,kf. (2)
73. (rogue adj3 periodical?).tw,kw,kf. (0)
74. (rogue adj3 publication?).tw,kw,kf. (0)
75. (rogue adj3 publish*).tw,kw,kf. (4)
76. (scam* adj3 edit*).tw,kw,kf. (3)
77. (scam* adj3 journal*).tw,kw,kf. (9)
78. (scam* adj3 periodical?).tw,kw,kf. (0)
79. (scam* adj3 publication?).tw,kw,kf. (0)
80. (scam* adj3 publish*).tw,kw,kf. (5)
81. (sham adj3 edit*).tw,kw,kf. (1)
82. (sham adj3 journal*).tw,kw,kf. (9)
83. (sham adj3 periodical?).tw,kw,kf. (0)
84. (sham adj3 publication?).tw,kw,kf. (1)
85. (sham adj3 publish*).tw,kw,kf. (50)
86. (spam* adj3 edit*).tw,kw,kf. (1)
87. (spam* adj3 journal*).tw,kw,kf. (4)
88. (spam* adj3 periodical?).tw,kw,kf. (0)
89. (spam* adj3 publication?).tw,kw,kf. (2)
90. (spam* adj3 publish*).tw,kw,kf. (5)
91. (unethic* adj3 edit*).tw,kw,kf. (21)
92. (unethic* adj3 journal*).tw,kw,kf. (22)
93. (unethic* adj3 periodical?).tw,kw,kf. (0)
94. (unethic* adj3 publication?).tw,kw,kf. (52)
95. (unethic* adj3 publish*).tw,kw,kf. (51)
96. (unprofessional* adj3 edit*).tw,kw,kf. (1)
97. (unprofessional* adj3 journal*).tw,kw,kf. (4)
98. (unprofessional* adj3 periodical*).tw,kw,kf. (0)
99. (unprofessional* adj3 publication?).tw,kw,kf. (3)
100. (unprofessional* adj3 publish*).tw,kw,kf. (1)
101. (untrust* adj3 edit*).tw,kw,kf. (0)
102. (untrust* adj3 journal*).tw,kw,kf. (0)
103. (untrust* adj3 periodical?).tw,kw,kf. (0)
104. (untrust* adj3 publication?).tw,kw,kf. (1)
105. (untrust* adj3 publish*).tw,kw,kf. (2)
106. pseudo-journal*.tw,kw,kf. (13)
107. pseudo-periodical*.tw,kw,kf. (5)
108. pseudo-publish*.tw,kw,kf. (2)
109. Beall* list.tw,kw,kf. (44)
110. 110 or/1-109 (1553)
111. limit 110 to yr=“2012-current” (1101)
112. Checklist/ use emczd,medall (24630)
113. Check Lists/ use eric (6639)
114. Editorial Policies/ use medall (7197)
115. guideline.pt. (16004)
116. Guidelines/ use eric (23366)
117. Guides/ use eric (8271)
118. exp Journalism/st use medall (998)
119. Open Access Publishing/st use medall (36)
120. exp Peer Review/st use medall (2244)
121. Publishing/st use medall (5272)
122. checklist*.tw,kw,kf. (117991)
123. check list*.tw,kw,kf. (19096)
124. guide*.tw,kw,kf. (1713265)
125. guidance*.tw,kw,kf. (319738)
126. criteria.tw,kw,kf. (1516630)
127. criterion.tw,kw,kf. (228194)
128. (tool or tools).tw,kw,kf. (1634450)
129. (instrument or instruments).tw,kw,kf. (564019)
130. algorithm?.tw,kw,kf. (504760)
131. instruction?.tw,kf,kw. (545248)
132. (inventory or inventories).tw,kf,kw. (308435)
133. (list or lists or listing or listings).tw,kf,kw. (461420)
134. 134 primer?.tw,kw,kf. (204194)
135. 135 or/112-134 [CHECKLISTS] (6958828)
136. 111 and 135 [PREDATORY JNL CHECKLISTS] (341)
137. (comment or editorial or news or newspaper article).pt. (1850264)
138. 136 not 137 [OPINION PIECES REMOVED] (286)
139. remove duplicates from 138 (206)

***************************

#### Peer review assessment: this section to be filled in by the reviewer

Reviewer: Kaitryn Campbell Date completed: 10 Nov 2018

Do you wish to be acknowledged? (If yes, the review team will be advised to add an acknowledgement to any publications related to this work). No – unless your organization requires it

The suggested acknowledgement is “We thank Xxxxx Yyyyyy, MLIS, AHIP (xxxxx Health Sciences Library, University of xxxxxx) for peer review of the MEDLINE search strategy.” [please edit to indicate your name, postnomials and institutional affiliation as you would like them presented].

**1. TRANSLATION**

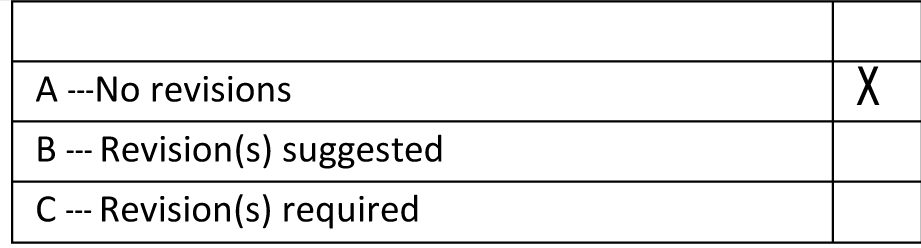

If “B” or “C,” please provide an explanation or example:

**2. BOOLEAN** and proximity **OPERATORS**

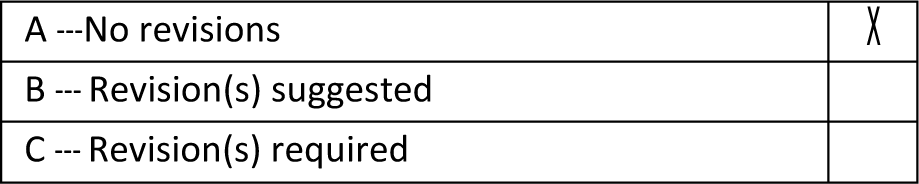

If “B” or “C,” please provide an explanation or example:

**3. SUBJECT HEADINGS**

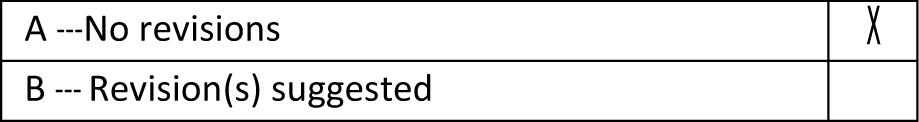

|  |
| --- |
| C --- Revision(s) required |

If “B” or “C,” please provide an explanation or example:

**4. TEXT WORD SEARCHING**

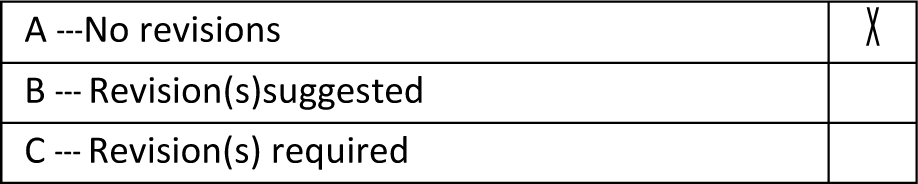

If “B” or “C,” please provide an explanation or example:

**5. SPELLING, SYNTAX, AND LINE NUMBERS**

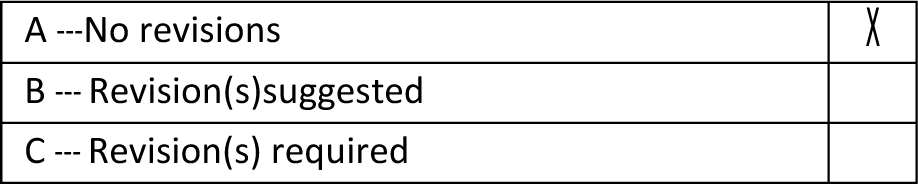

If “B” or “C,” please provide an explanation or example:

6. **LIMITS AND FILTERS**

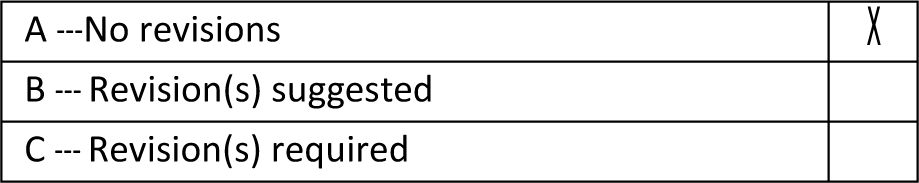

If “B” or “C,” please provide an explanation or example:

OVERALL EVALUATION (Note: If one or more “revision requried” is noted above, the response below must be “revisions requried”)

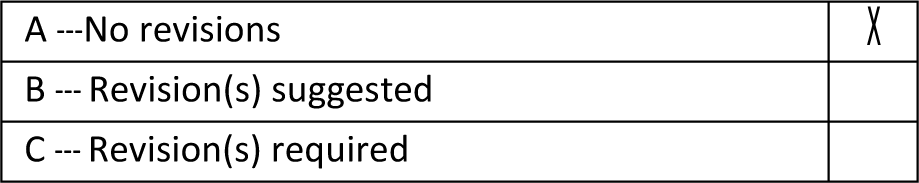

Additional comments:

Solidly done. No errors or omissions found. Re: including just the “predatory journals” concept alone as the strategy for the bibliographic database searching, this seems like a strong option for 2 reasons: 1) limited retrieval numbers; 2) I have some experience doing “checklist” searches and found the description of these types of items can be extremely variable. My personal preference would be to leave the checklist concept out.

## Appendix 2 Ovid Multifile

Database: Embase Classic+Embase <1947 to 2018 November 19>, Ovid MEDLINE(R) ALL <1946 to November 19, 2018>, PsycINFO <1806 to November Week 2 2018>, ERIC <1965 to October 2018>

Search Strategy:

--------------------------------------------------------------------------------

1. (predator* adj3 edit*).tw,kw,kf. (29)
2. (predator* adj3 journal*).tw,kw,kf. (397)
3. (predator* adj3 periodical?).tw,kw,kf. (6)
4. (predator* adj3 publication?).tw,kw,kf. (49)
5. (predator* adj3 publish*).tw,kw,kf. (379)
6. (bogus adj3 edit*).tw,kw,kf. (2)
7. (bogus adj3 journal*).tw,kw,kf. (7)
8. (bogus adj3 periodical?).tw,kw,kf. (0)
9. (bogus adj3 publication?).tw,kw,kf. (0)
10. (bogus adj3 publish*).tw,kw,kf. (1)
11. (dark adj3 edit*).tw,kw,kf. (32)
12. (dark adj3 journal*).tw,kw,kf. (9)
13. (dark adj3 periodical?).tw,kw,kf. (4)
14. (dark adj3 publication?).tw,kw,kf. (2)
15. (dark adj3 publish*).tw,kw,kf. (19)
16. (decepti* adj3 edit*).tw,kw,kf. (21)
17. (decepti* adj3 journal*).tw,kw,kf. (15)
18. (decepti* adj3 periodical?).tw,kw,kf. (0)
19. (decepti* adj3 publication?).tw,kw,kf. (3)
20. (decepti* adj3 publish*).tw,kw,kf. (20)
21. (disreput* adj3 edit*).tw,kw,kf. (0)
22. (disreput* adj3 journal*).tw,kw,kf. (3)
23. (disreput* adj3 periodical?).tw,kw,kf. (0)
24. (disreput* adj3 publication?).tw,kw,kf. (3)
25. (disreput* adj3 publish*).tw,kw,kf. (0)
26. (distrust* adj3 edit*).tw,kw,kf. (1)
27. (distrust* adj3 journal*).tw,kw,kf. (2)
28. (distrust* adj3 periodical?).tw,kw,kf. (0)
29. (distrust* adj3 publication?).tw,kw,kf. (0)
30. (distrust* adj3 publish*).tw,kw,kf. (5)
31. (exploit* adj3 edit*).tw,kw,kf. (107)
32. (exploit* adj3 journal*).tw,kw,kf. (29)
33. (exploit* adj3 periodical?).tw,kw,kf. (1)
34. (exploit* adj3 publication?).tw,kw,kf. (37)
35. (exploit* adj3 publish*).tw,kw,kf. (96)
36. (fake? adj3 edit*).tw,kw,kf. (11)
37. (fake? adj3 journal*).tw,kw,kf. (37)
38. (fake? adj3 periodical?).tw,kw,kf. (0)
39. (fake? adj3 publication?).tw,kw,kf. (4)
40. (fake? adj3 publish*).tw,kw,kf. (20)
41. (hoax$2 adj3 edit*).tw,kw,kf. (1)
42. (hoax$2 adj3 journal*).tw,kw,kf. (5)
43. (hoax$2 adj3 periodical?).tw,kw,kf. (0)
44. (hoax$2 adj3 publication?).tw,kw,kf. (2)
45. (hoax$2 adj3 publish*).tw,kw,kf. (4)
46. (illegitim* adj3 edit*).tw,kw,kf. (3)
47. (illegitim* adj3 journal*).tw,kw,kf. (19)
48. (illegitim* adj3 periodical?).tw,kw,kf. (0)
49. (illegitim* adj3 publication?).tw,kw,kf. (6)
50. (illegitim* adj3 publish*).tw,kw,kf. (12)
51. (mislead* adj3 edit*).tw,kw,kf. (42)
52. (mislead* adj3 journal*).tw,kw,kf. (36)
53. (mislead* adj periodical?).tw,kw,kf. (0)
54. (mislead* adj3 publication?).tw,kw,kf. (57)
55. (mislead* adj publish*).tw,kw,kf. (5)
56. (non-legitim* adj3 edit*).tw,kw,kf. (0)
57. (non-legitim* adj3 journal*).tw,kw,kf. (0)
58. (non-legitim* adj3 periodical?).tw,kw,kf. (0)
59. (non-legitim* adj3 publication?).tw,kw,kf. (0)
60. (non-legitim* adj3 publish*).tw,kw,kf. (0)
61. (questionabl* adj3 edit*).tw,kw,kf. (24)
62. (questionabl* adj3 journal*).tw,kw,kf. (38)
63. (quesionabl* adj3 periodical?).tw,kw,kf. (0)
64. (questionabl* adj3 publication?).tw,kw,kf. (44)
65. (questionabl* adj3 publish*).tw,kw,kf. (48)
66. (racket? adj3 edit*).tw,kw,kf. (0)
67. (racket? adj3 journal*).tw,kw,kf. (1)
68. (racket? adj3 periodical?).tw,kw,kf. (0)
69. (racket? adj3 publication?).tw,kw,kf. (0)
70. (racket? adj3 publish*).tw,kw,kf. (0)
71. (rogue adj3 edit*).tw,kw,kf. (4)
72. (rogue adj3 journal*).tw,kw,kf. (2)
73. (rogue adj3 periodical?).tw,kw,kf. (0)
74. (rogue adj3 publication?).tw,kw,kf. (0)
75. (rogue adj3 publish*).tw,kw,kf. (4)
76. (scam* adj3 edit*).tw,kw,kf. (3)
77. (scam* adj3 journal*).tw,kw,kf. (9)
78. (scam* adj3 periodical?).tw,kw,kf. (0)
79. (scam* adj3 publication?).tw,kw,kf. (0)
80. (scam* adj3 publish*).tw,kw,kf. (6)
81. (sham adj3 edit*).tw,kw,kf. (1)
82. (sham adj3 journal*).tw,kw,kf. (9)
83. (sham adj3 periodical?).tw,kw,kf. (0)
84. (sham adj3 publication?).tw,kw,kf. (1)
85. (sham adj3 publish*).tw,kw,kf. (50)
86. (spam* adj3 edit*).tw,kw,kf. (1)
87. (spam* adj3 journal*).tw,kw,kf. (4)
88. (spam* adj3 periodical?).tw,kw,kf. (0)
89. (spam* adj3 publication?).tw,kw,kf. (2)
90. (spam* adj3 publish*).tw,kw,kf. (5)
91. (unethic* adj3 edit*).tw,kw,kf. (21)
92. (unethic* adj3 journal*).tw,kw,kf. (22)
93. (unethic* adj3 periodical?).tw,kw,kf. (0)
94. (unethic* adj3 publication?).tw,kw,kf. (52)
95. (unethic* adj3 publish*).tw,kw,kf. (51)
96. (unprofessional* adj3 edit*).tw,kw,kf. (1)
97. (unprofessional* adj3 journal*).tw,kw,kf. (4)
98. (unprofessional* adj3 periodical*).tw,kw,kf. (0)
99. (unprofessional* adj3 publication?).tw,kw,kf. (3)
100. (unprofessional* adj3 publish*).tw,kw,kf. (1)
101. (untrust* adj3 edit*).tw,kw,kf. (0)
102. (untrust* adj3 journal*).tw,kw,kf. (0)
103. (untrust* adj3 periodical?).tw,kw,kf. (0)
104. (untrust* adj3 publication?).tw,kw,kf. (1)
105. (untrust* adj3 publish*).tw,kw,kf. (2)
106. pseudo-journal*.tw,kw,kf. (13)
107. pseudo-periodical*.tw,kw,kf. (5)
108. pseudo-publish*.tw,kw,kf. (2)
109. Beall* list.tw,kw,kf. (46)
110. or/1-109 (1564)
111. 111 limit 110 to yr=“2012-current” (1112)
112. (comment or editorial or news or newspaper article).pt. (1853727)
113. 111 not 112 [OPINION PIECES REMOVED] (869)
114. remove duplicates from 113 (586)
115. 114 use medall (333)
116. 114 use emczd (156)
117. 114 use eric (16)
118. 18 114 not (115 or 116 or 117114 not (115 or 116 or 117) (81)

